# Current gaps in survey design and analysis for molecular xenomonitoring of vector-borne neglected tropical diseases: a systematic review

**DOI:** 10.1101/2025.01.16.25320642

**Authors:** Angus McLure, Tilahun Alamnia, Zhiwei Xu, Colleen L. Lau, Helen J. Mayfield

## Abstract

**Objectives:** Molecular Xenomonitoring (MX) is a surveillance method for vector-borne diseases where vectors are tested for molecular pathogen markers. Testing is typically on pools (groups) of vectors. MX is a sensitive and efficient complement to human based surveillance. However, there is limited guidance about the appropriate design and analysis of MX surveys. We reviewed the literature to understand the common objectives, survey designs, and analysis methods for MX surveys for two vector-borne neglected tropical diseases: lymphatic filariasis (LF) and onchocerciasis.

**Methods:** We searched peer-reviewed literature for studies published between 1999 and 2022 that presented the results of surveys that collected vectors in field surveys and used a molecular test for the presence of the causative pathogens for LF and onchocerciasis.

**Results:** A total of 76 studies (LF: 45; onchocerciasis: 31) across 30 countries were included in the review. The five most common objectives were determination of elimination status after mass drug administration, comparison of vector and human infection indicators, evaluation of an intervention, comparison of vector collection methods, and comparison of laboratory techniques. Nearly all studies used a cluster or hierarchical sampling frame to collect vectors (72/76), but very few studies accounted for this in their designs (2/76) or analysis (1/76). While few studies justified the number of vectors included in each pool (5/76), nearly all studies accounted for pooled testing when calculating pathogen prevalence from results (69/76). Few studies justified the number or selection of sampling sites or total sample size (16/76).

**Conclusions:** Published MX surveys for LF and onchocerciasis had varied objectives, study designs and analysis methods, but proper consideration of survey design was frequently missing from the analysis. There is a need for statistical tools and guidance to enable appropriate design and analysis of MX surveys while accounting for disease, objective, and context-specific considerations.

## Introduction

Neglected Tropical Diseases (NTDs) remain a major cause of mortality and morbidity, disproportionality affecting low-income countries [1]. Several of these diseases are the focus of targeted global elimination campaigns that rely on mass drug administrations as the primary intervention [2]. For these programs to meet their objectives, efficient and effective surveillance tools that can detect evidence of infection or transmission in specific populations and locations are crucial to facilitate informed, evidence-based decision making. This requirement is particularly critical as these programmes approach their end game, when the prevalence of the diseases become very low and often more focal [3–7]. Molecular xenomonitoring (MX), the collection of disease vectors and other biting invertebrates and testing them for the presence of molecular markers of the pathogen, is one strategy that can provide evidence of pathogen presence and transmission potential in an area [8].

MX studies can support several specific objectives of disease surveillance, including detection of the pathogen signals (or establish absence) in a study area [9–15], comparing the prevalence of the pathogen marker in the vectors to a threshold to inform programmatic decisions [5, 16–18], or comparing the prevalence of the pathogen marker in two samples of vectors, e.g. before and after an intervention [19–23]. However, before MX can be applied, initial studies will need to focus on validating the tool and identifying effective ways of capturing and testing the vectors [24–27]. A key part of this validation is an understanding of the most appropriate survey design and how best to analyse the results [28, 29]. Once the use of MX has been validated for the pathogen and vector it can be used as a standalone method, or in conjunction with human surveillance.

MX is often used as a complement to disease surveillance in humans [9, 10, 12, 13, 21] and has its own set of advantages and disadvantages. MX can be less intrusive than human surveillance methods which often involve taking blood or tissue samples. Though highly dependent on local conditions including weather and climate, it is often possible to sample very large numbers of vectors, far more than the number of humans who could be tested for a similar amount of survey effort [21, 25] and therefore potentially resulting in a higher probability of detecting a signal where the prevalence is targeted markers comparable in humans and vectors. To reduce time and costs, vectors are often tested in pools (groups) rather than individually, with a single positive or negative test result for the whole pool. Though not an issue for surveys attempting to establish the presence or absence of a pathogen in a vector population, pooled testing leads to a loss of information when it comes to estimate the prevalence of the pathogen [30].

There are several characteristics of MX surveys that complicate their design and analysis. In addition to the pooled testing methodology, MX surveys often utilise cluster designs or hierarchical sampling frame, with the vectors collected at a number of collection sites across the area of interest. Both cluster sampling and pool testing can reduce the total cost or effort of the survey but reduce the effective sample size and complicate design and analysis [31].

Efficient and appropriate survey designs and analysis plans are therefore important to maximise the information gained for minimal cost. Early consideration of design and analysis is critical to provide the best evidence to support decision making in disease elimination programmes. However, there is relatively little published research that assesses the appropriateness of common MX survey designs. Practical advice on key elements of survey design is also lacking. Understanding the common objectives, design, and analysis of MX surveys is the key first step before this gap can be addressed.

There are several existing reviews of MX for NTDs. Pilotte et al. [8] provide an excellent overview of the methods, strengths, and operational research gaps for MX for two diseases: onchocerciasis, transmitted by biting blackflies of the *Simulium damnosum* complex; and lymphatic filariasis (LF), transmitted by mosquitos of the genera *Aedes, Culex, Anopheles,* and *Mansonia*. Pryce and Reimer [32] and Pryce et al. [33] evaluated the sensitivity of MX surveys to detect locations of people with microfilaremia. Reimer and Pryce [34] examined the effect of vector sampling methods and vector genus on prevalence of mosquitos positive for filarial DNA.

In this systematic review, we also focus on onchocerciasis and LF. While MX has been used extensively for both diseases, it is part of the package of activities that the WHO requires to certify elimination status for onchocerciasis [35], but not LF. Unlike previous reviews, we take a statistical perspective on MX survey design and analysis. We ask the following questions of MX surveys published in peer-reviewed literature:

- What were the *objectives* of MX surveys?
- What survey *designs* (e.g. site selection, sample sizes, pooling strategies) were used for MX and how are these designs selected?
- How were data from MX surveys *analysed*?
- Were the above considerations (objective, design, and analysis) well aligned and how could this alignment be improved?

## Methods

An initial literature search was performed on PubMed database for articles published between January 1999 and September 2022 (date of search). The criteria for inclusion were titles or abstracts that included a term related to MX (molecular xenomonitoring OR mosquito surveillance OR vector surveillance) as well as a term related to one of the target diseases (lymphatic filariasis OR onchocerciasis). Titles were imported into Covidence for screening [36]. Title and abstract screening were conducted by two reviewers (TA and ZX) with conflicts reviewed by discussion and consensus. Full text screening was conducted by two reviewers (TA and AM).

The following types of studies were excluded: reviews without new analysis/presentation of data; studies that did not collect vectors or other biting insects (e.g. if only humans were sampled); studies where no insects were tested using a molecular test for the presence of the pathogen; and studies that did not present data from a field survey (e.g. simulation studies; studies to validate molecular testing on experimentally infected vectors); articles not in English. The following types of studies were *not* excluded unless they also met one of the above exclusion criteria: reanalysis/secondary analysis of data; studies that tested insects but found all to be negative for the pathogen marker; studies where some vectors were tested individually (rather than in pools).

Data extraction was conducted by two reviewers (HM and AM) and included the following variables: year of publication; disease (LF or onchocerciasis); country(ies) or territory(ies) where insects were collected; study objective(s); whether the study used a hierarchical survey design and details including site selection; total number of insects caught and/or tested; justification of the sample size; number of pools tested; pooling strategy and justification for this strategy; whether insect species were separated before pooling and testing; software used to analyse survey data; whether analysis of data was hierarchical and details including the levels at which the analysis was conducted.

To help classify studies with complex or multiple objectives, each study was classified as addressing one or more of the following: validation of elimination following an MDA program; evaluation of an intervention with pre and post surveys; comparison of MX indicators to human-based indicators; comparison of laboratory techniques for detecting pathogen DNA in mosquitos; and comparison of vector collection methods. Where appropriate, extracted results were summarised using counts and percentages in R [37].

## Results

### Included studies by country and disease

The database search identified 1225 unique studies. Of these, 1120 studies were excluded based on abstract and title. Of the 105 remaining studies, one was excluded as we could not retrieve the full text, two studies were excluded because they were not on LF or onchocerciases, three studies were excluded because no insects were caught, 14 were excluded because they did not use a molecular test to detect pathogen DNA in insects, and nine were excluded because they did not include insects from field surveys (Fig. 1). A total of 76 studies were included in the review: 31 on onchocerciasis [5, 12, 13, 16, 18, 23, 38–62] and 45 on LF [7, 9–11, 14, 15, 17, 19–22, 24–29, 63–90]. No study covered both LF and onchocerciasis.

**Fig. 1:**
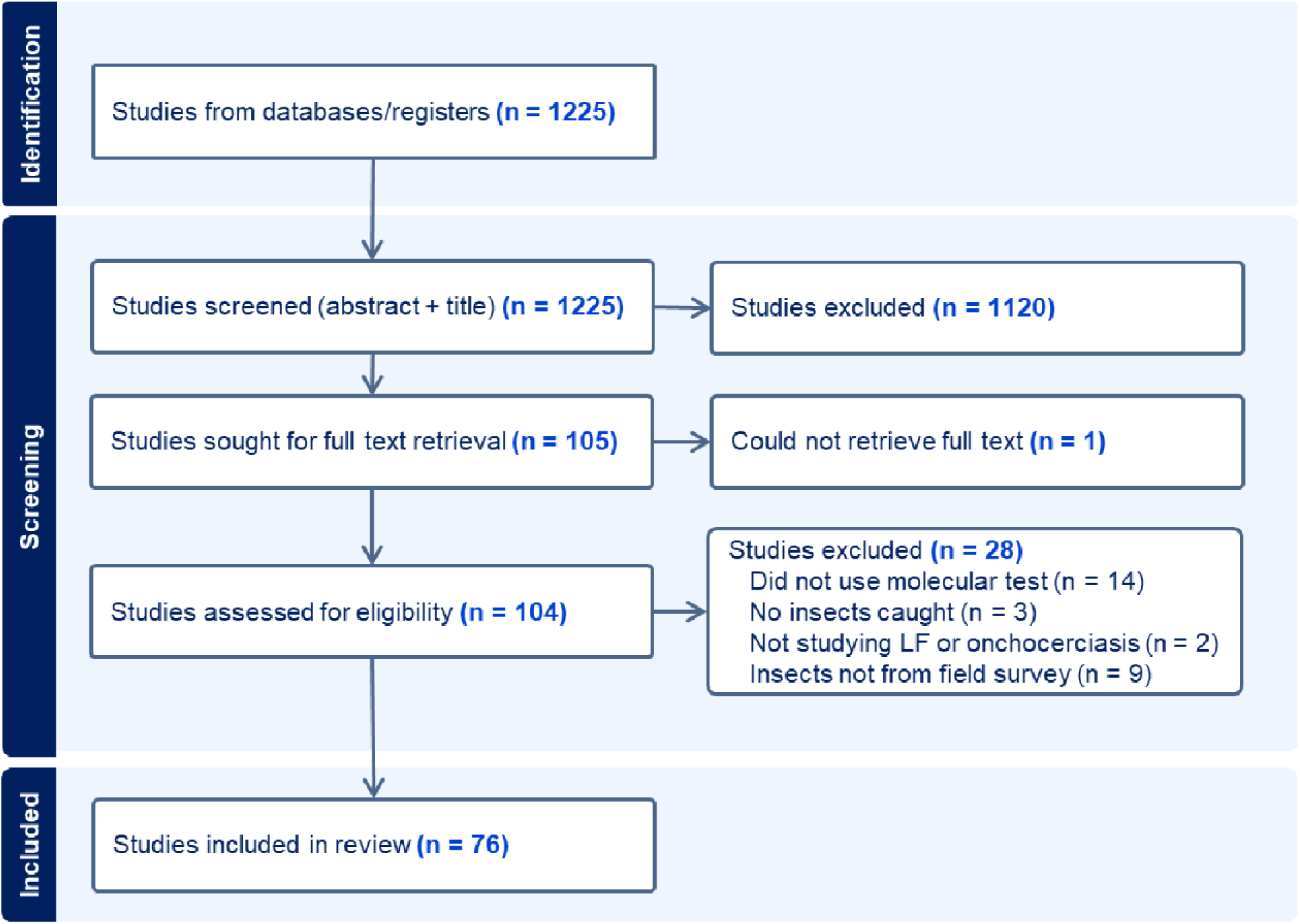
Summary of number of papers identified and screened, including reasons for exclusion.

The included studies were conducted across 30 countries, with LF studies in 20 countries and onchocerciasis studies in 15 countries. Five studies were undertaken across more than one country and five countries had studies for both diseases. Sri Lanka had the most studies for LF (seven) and Mexico had the most studies for onchocerciasis (seven). Nigeria had five studies; one for LF and four for onchocerciasis. By region, the largest number of studies were conducted in Africa (20 LF and 15 onchocerciasis). The remaining 16 studies of onchocerciasis were all in the Americas, while the remaining studies of LF were distributed across Asia (14) and the Pacific (8), with three studies in the Americas (Brazil).

### Study objectives

Studies had a wide range of objectives. The most common objective overall (48/76 studies) and for onchocerciasis studies (27/31 studies) was determination of elimination status (i.e. validation) post-MDA. Most studies (LF: 27/45; onchocerciasis: 18/31; overall: 45/76) also made comparisons between indicators of infection markers in humans (e.g. detection of microfilaria or antigens) and MX indicators. One quarter of studies (19/76) evaluated interventions by comparing MX indicators in surveys before and after the intervention(s). Thirteen studies, all for LF, compared different insect collection techniques. Eighteen studies, predominantly on LF (13), compared different lab techniques for detecting pathogen DNA in the insects. The number of studies addressing each of the five most common objectives are listed in Fig. 2.

**Fig. 2:**
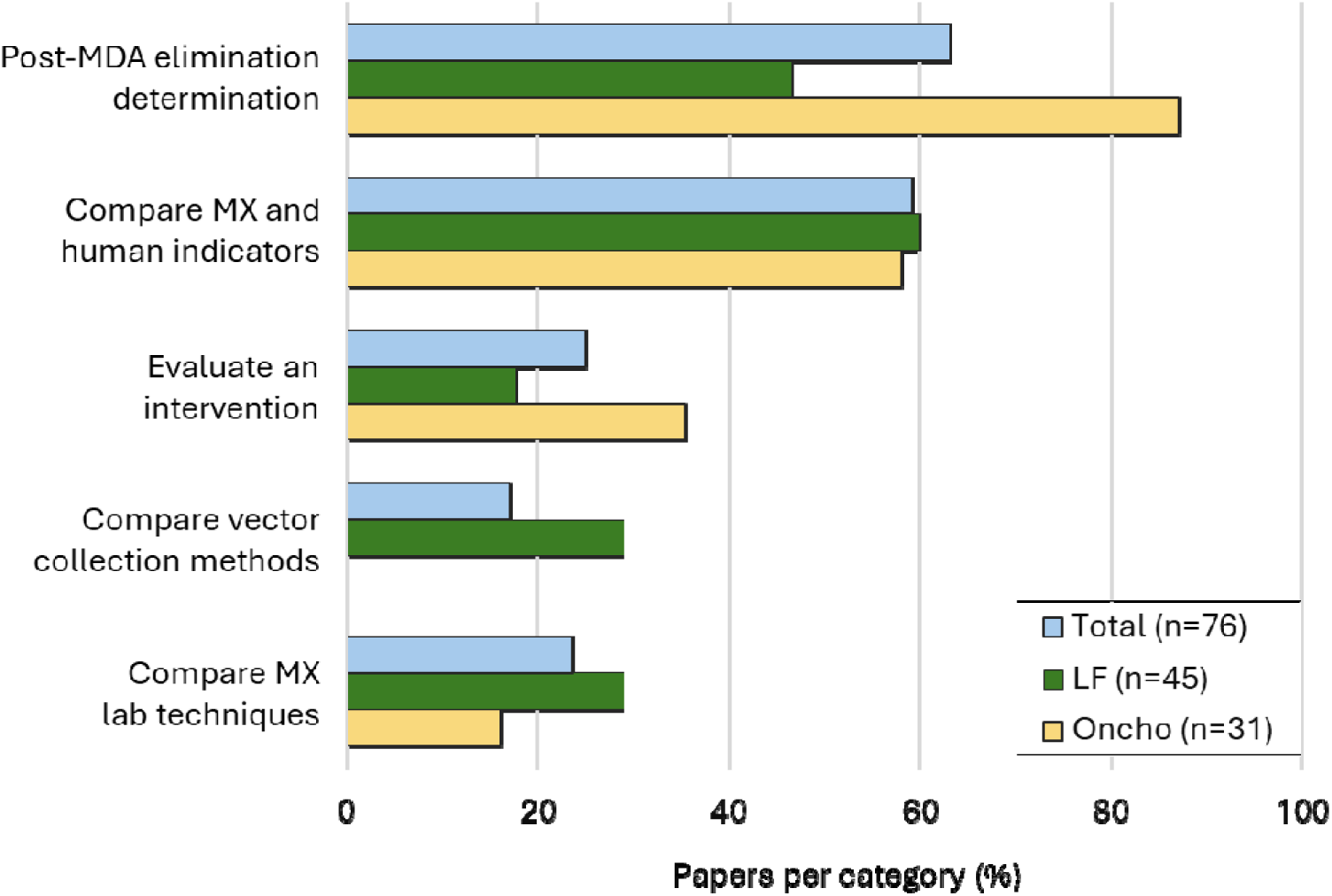
Frequency of the five most common objectives of MX surveys in the included studies by disease and overall. Some studies had multiple objectives. Values are provided in Supplementary Table S1. Abbreviations LF: lymphatic filariasis; Oncho: onchocerciasis.

#### Post-MDA elimination determination

Nearly all studies of onchocerciasis (27/31) and half of LF studies (21/45) aimed to evaluate progress towards elimination in settings where multiple rounds of MDA had been conducted. Many of these onchocerciasis studies compared the proportion of infective and infected blackflies to thresholds set by the WHO [35, 91] to validate elimination status and evaluate need for further MDA. Most onchocerciasis studies with this objective confirmed that prevalence had reached very low levels, with about half (14/27) reporting no detections of positive blackflies, and a number of studies with longitudinal sampling reporting no detections in the latter years [16, 57]. For LF, where there are only provisional WHO targets for thresholds to inform decisions to stop or start MDA using MX markers [92], several studies have proposed alternative provisional thresholds for various vector genera [22, 82, 93]. These values have then been used in a number of subsequent studies that compared prevalence to one of these thresholds [7, 25, 28]. Some LF studies evaluating progress towards elimination in post-MDA settings reported no detection of filarial DNA in mosquitoes (6/21 studies).

#### Comparing MX and human indicators

MX studies were often conducted alongside or compared to results of surveillance of human indicators in the same geographical area: 18/31 (58%) onchocerciasis studies and 27/45 (60%) of LF studies. In these studies, human participants would be screened for indicators of current or past infection including filarial antigens, anti-filarial antibodies, and detection of microfilaria in skin or blood using microscopy. For many of these studies, particularly onchocerciasis studies in which the primary objective was to evaluate progress towards elimination, the primary comparison was between the indicators (human and entomological) and respective thresholds with the two types of indicators providing independent lines of evidence for or against elimination. However, in a number of LF studies, human and entomological indicators were compared in terms of sensitivity to detect low prevalence of pathogen markers [21, 25, 29, 74, 82, 83]. Some studies also compared the cost of human surveillance and MX, such as Subramanian et al [25] that found that MX had a similar cost but was more sensitive for detecting signals LF in a population than standard Transmission Assessment Surveys (TAS) in children.

#### Evaluating an intervention

Of the studies that evaluated an intervention (19/76), all but one evaluated MDA, with a single study evaluating the effect of bed-nets [19]. In some studies, the intervention (MDA) was conducted repeatedly with surveys before, between, and after rounds. In these studies, the prevalence of filarial markers in insects or other entomological markers were compared between repeated surveys. Most studies evaluating an intervention (LF: 7/8; Onchocerciasis: 7/11) used MX alongside human indicators. Studies overwhelmingly found that when there was any trend between pre- and post-surveys, human and MX indicators showed the same trend. Notably, McPherson et al. [21] reported that within a year of the intervention (MDA), prevalence of filarial DNA had significantly declined, but did not detect a significant decline in filarial antigen prevalence in humans.

#### Comparing vector collection methods or lab techniques

All studies comparing vector collection methods came from the LF literature and mostly focused on the yields of mosquitos caught by different types of traps or human landing catches. One study [29] considered the number of trapping locations, comparing the estimates of filarial DNA prevalence when conducting intensive sampling at a few sites versus collection of the same number of mosquitos at a larger number of sites. Studies comparing laboratory techniques were also mostly of LF (13/18), and usually compared a molecular test such as PCR detection of filarial DNA to dissection of insects for microscopy. However, a number of studies compared molecular techniques to each other, e.g. comparing real-time PCR, LAMP, and dissection [23, 79]; comparing simple and multiplex PCR [66]; or comparing novel high-throughput automated PCR systems to existing PCR methods [58]. The primary comparison was that of sensitivity to detect the pathogen, with studies concluding that molecular techniques were as sensitive [23, 53] or more sensitive [63, 65, 89] than microscopy. Two studies also compared the cost of detection methods, either dissection vs PCR [61], or dissection vs PCR vs PCR-ELISA [89].

Other less common objectives not included in Fig. 2 were also found. Several studies used MX to identify or exclude the possibility of transmission in areas not previously known to be endemic and had no history of MDA [14, 38, 72, 73, 78]. Some studies, primarily of onchocerciasis, reported longitudinal post-MDA surveillance attempting to detect signs of recrudescence or establish that prevalence continued to decline after cessation of MDA [9, 39, 48, 54, 56, 57, 59, 60, 82, 83]. Many studies measured entomological indices beyond the detection of pathogen DNA Annual biting rates were sometimes calculated in LF studies [79] and often calculated for onchocerciasis studies [12, 16, 43, 47, 51, 57, 59] as an intermediary to calculating the number of infective bites per person per year. Other studies focused on detailed analyses of the composition of vector species present [77], or compared vectorial capacity between present species [41], or measured the prevalence of insecticide resistance [77], or vector dispersion using mark-release-recapture experiments [24]. Other studies tried to understand the environmental factors influencing vector abundance in a region [23] or around households [75]. Other studies compared variability of vector abundance, biting rates, or prevalence of filarial DNA across different seasons [12, 23, 80], with one study attempting to identify correlation between environmental factors (rainfall), vector abundance, and prevalence of filarial DNA in vectors [85]. A few studies reported the total and itemised costs for MX surveys [11, 25].

### Survey designs

#### Hierarchical sampling frame

The included studies employed a wide variety of survey designs to collect, pool, and test disease vectors. Nearly all the surveys (onchocerciasis: 29/31; LF: 43/45) utilised a cluster or hierarchical sampling frame, with the sampling frame being unclear for three studies [61, 64, 66], and one study collecting insects from a single site [23] (Fig. 3). In one study, vectors were collected from a single site in each study area [38]. However, the majority of studies had more complex, nested sampling frames. In some studies, a number of study areas were selected non-randomly (e.g. villages, rivers, cities), with more than one collection site (traps or human landing catches) in each study area of interest [12, 14, 22, 39, 79, 80, 87, 90]. Other studies used multi-stage cluster sampling, randomly or systematically selecting geographical units (e.g. village or a small public health unit) from the survey area(s), and then selecting multiple vector collection sites in each geographical unit [28, 29, 67, 82, 86].

**Fig. 3:**
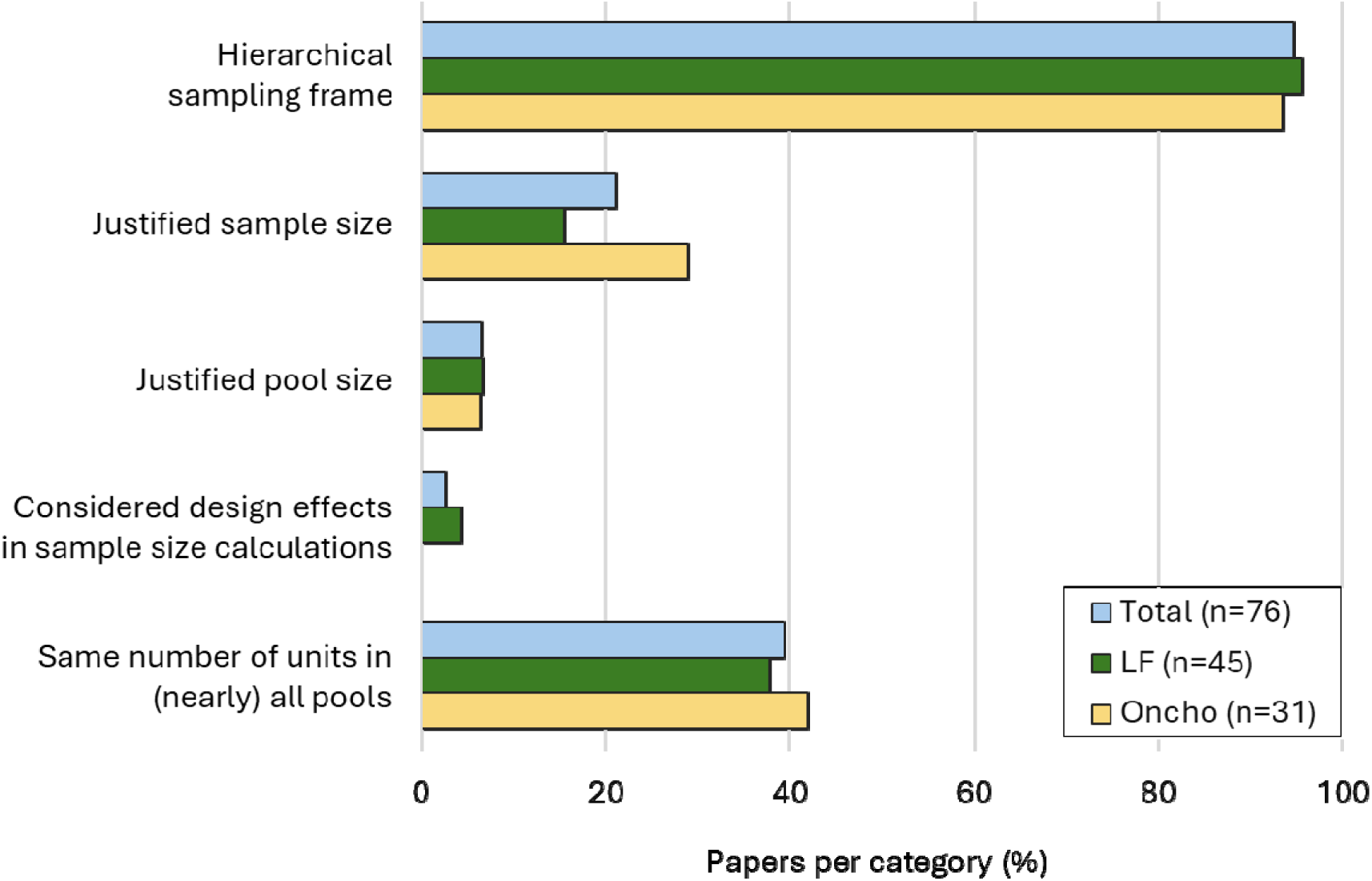
Frequency of key design choices and justifications for the MX surveys in the included studies by disease and overall. Values are provided in Supplementary Table S2. Abbreviations LF: lymphatic filariasis; Oncho: onchocerciasis.

#### Site selection

The selection of collection sites was randomised or systematic in some studies [7, 17, 21, 75], and purposive in others, usually targeting high risk locations where infected humans or vectors had been identified previously [10, 50, 52, 65, 79], but sometimes based on accessibility [51]. Some surveys adopted multi-stage sampling using a mix of purposive and randomised selection at the different sampling levels. For example Derua et al. (2017) purposively selected villages and hamlets within villages to maximise ease of access and mosquito collections, but three households were randomly selected from each hamlet for vector collection [65].

More than half of the surveys conducted MX surveillance alongside surveillance in humans (onchocerciasis: 18/31; LF: 27/45) (Fig. 3). For onchocerciasis studies, vector collection was often conducted near breeding sites (river banks) rather than households [23, 38]. However, many MX surveys, especially LF surveys, were conducted in or near homes, including concurrent MX and human surveillance studies [9, 21, 81] and MX-only studies [28]. In a few studies, insect collection sites were chosen to be very close to the households of participants in the human surveys [21, 81]. In others, extensive sampling of households in both the human and entomological studies meant some household locations were included in both arms of the study [22, 72]. In most studies however the human and MX studies were co-located only at a higher level of spatial aggregation, e.g. the same villages or same public health units, but not the same households.

#### Sample size of insects

The number of insects collected was generally very large, but with substantial variability between included studies. In some studies, only a portion of all collected vectors were tested for filarial DNA using molecular methods and in some cases this portion was less than 30% of the total [65, 73] or as little as 2% [77]. The number of insects examined with a molecular test was the most reliably reported and readily comparable measure of sample size between studies, being reported in all but one study [71]. Using this metric, the sample size of insects was generally smaller for LF studies (median: 7,900; IQR: 3,100-15,000) than for onchocerciasis studies (median: 31,000, IQR: 12,000-86,000). Onchocerciasis studies where one of the objectives was post-MDA elimination determination generally had larger sample sizes than studies without this objective (median: 34,000, IQR: 15,000-97,000 vs median: 11,000, range: 7,500-13,000). LF studies that aimed to determine elimination status after MDA had similar sample sizes (median: 8,500; range: 4,000-23,000) to LF studies without this objective (median: 5,800; IQR: 2,500-15,000).

#### Pooling schemes

Pooling schemes varied substantially between studies. Studies stratified insect pools by one or more variables (e.g. collection site, collection time, collection method, vector species), though these were not always clearly specified. Stratification methods included pooling by collection site and time with the time interval sometimes as short as one hour [16, 38, 63] but often longer [20, 25, 28, 66, 81, 85, 87]; or pooling by village/community and method of collection but combining samples from different collection sites [11, 12, 39, 52, 55, 78, 89]; or with different pooling strategies in different study years [21, 41]. Some studies chose not to separate insects by species for some or all of their surveys [41] or separated only to genus level [17, 73, 74, 78, 79]. In many onchocerciasis studies, heads and bodies of flies were separated. In some studies, only pools of heads were tested [5, 18, 45], or bodies were tested first followed by testing heads only in locations with positive body pools [16, 54–56]. Though much less common, at least one LF study [70] divided mosquitoes into body segments to test heads separately from the thorax and abdomen.

The maximum number of insects per pool varied substantial between studies and was typically larger in onchocerciasis studies (median: 50, range: 20-300) than LF studies (median: 20, range: 1-30). In some studies, the number of units per pool was the same across all (or nearly all) pools, while in others a range of pools sizes were used. Of the onchocerciasis studies, 13 used fixed pool size, 14 a variable pool size and four studies were unclear. Of the LF studies, 17 used a fixed pool size, 23 a variable pool size and five studies were unclear. Similarly, some studies capped the number and size of pools per site [69], not testing any collected insects beyond these limits.

#### Justification of survey designs

Few studies provided any justification for the survey design (e.g. sampling frame, site selection, sample size, pooling scheme), with others providing justifications for only some of the design choices. No studies provided justification for the number of insects per pool on statistical grounds. One LF study [76] cited a WHO handbook [94] which states that “a pool of 25 mosquitoes is often used for PCR processing in determining infection”, and did not provide any other justification. Another LF study [79] stated that the choice of pool size was based on a previous study [95] that compared different pool sizes (range: 25 to 200), but used smaller pools in their own study (range: 5-20) without further justification. Two onchocerciasis studies validated the sensitivity and specificity of the molecular test using known positive and negative pools with a range of sizes before applying the largest verified pool size to the field survey component of their study [53, 58], however did not provide statistical justification for using the largest validated pool size.

Most studies (LF: 38/45; onchocerciasis: 22/31) provided no justification of sample sizes beyond trying to catch as many insects or vectors as possible (Fig. 3). The most common justification for sample size in onchocerciasis studies (six studies) was to collect enough blackflies such that if all were negative that one-sided 95% confidence interval for prevalence would be less than 0.05% [12, 13, 39, 42, 59, 60], citing guidelines published by the WHO; however, these studies reported different numbers of flies (3,900 or 6,000) needed to achieve the same goal. Six LF studies [11, 20, 21, 69, 78, 79] justified their choice of sample size in terms of power to determine whether prevalence was below a given threshold value and another LF study [28] set target sample sizes based on desired precision of prevalence estimate. Two LF studies [21, 69] included design effects in their sample size calculations to account for cluster sampling designs, and a third study [28] specifically stated that they didn’t include a design effect (i.e. design effect of 1); however, none of these studies justified their choice of design effects. No onchocerciasis studies discussed the inclusion of design effects.

Studies rarely discussed or justified the number of collection sites (clusters) on statistical grounds. A notable exception [29] compared different study designs with the same total sample size but different number of collection sites, finding that point estimates of prevalence were similar across designs.

### Data analysis

Nearly all studies used an analysis method that could estimate insect-level prevalence from the pooled data (onchocerciasis: 30/31; LF: 39/45). In studies where all pools were negative (onchocerciasis: 15/31; LF: 9/45), analysis did not require specialised software to adjust for pooled testing. Most studies used the Poolscreen software [96] to make the appropriate adjustment for the pooled testing protocol (onchocerciasis: 30/31; LF: 28/45). Two studies used R packages: Takagi et al. [85] used binGroup [97], and McPherson et al. [21] used PoolTestR [98]. Two studies [47, 66] had a fixed pool size and a further two studies [19, 67] appear to have tested insects individually, in which case prevalence could be estimated with a simple formula and did not require specialised software. Three studies with positive insect pools [14, 27, 88] only reported pool-level results, or did not clearly state whether the results were adjusted for pooling. While most studies reported confidence intervals for estimates of prevalence, some studies did not [23, 24, 47, 65–68, 88, 89], especially where there were no positives insects detected in the study [15, 24, 64, 72, 76, 77] or subpopulation [70, 86]. While nearly all studies used a hierarchical sampling design and most studies estimated prevalence in an area by aggregating the test results from pools from multiple sites, only one study [21] adjusted prevalence estimates for clustering at sampling sites, using the PoolTestR R package [99].

Many studies compared the prevalence of pathogen markers in insects from two or more samples. The samples could be from different areas [21, 28, 41, 44, 75], different timepoints in the same areas [7, 9, 12, 16, 23, 39, 49, 57, 59, 83, 87], different insect species [21, 41], different trapping methods [75], or different detection methods [63, 86, 89]. Some studies used common statistical tests to examine the difference in the proportion of positive pools between samples, such as the chi-squared [28], Fisher’s exact [53, 63], Kruskal-Wallis [23], and t-tests [75]. In some of these studies, there were different numbers of insects in each pool, sometimes with systematic differences between samples [7, 28, 63, 83], and as none of these tests account for pool size, differences in pool sizes between samples may have masked or exaggerated any true difference in insect-level prevalence between samples.

In some studies, the confidence intervals around estimates were non-overlapping and the samples could be considered independent, and therefore no further test was necessary to establish a difference between samples [44, 86]. However, even in some studies where a primary objective was to determine prevalence difference between two samples (e.g. before and after an intervention or different detection methods), there was often no quantification of the difference between the samples (e.g. prevalence ratios, prevalence differences), or no confidence interval for this difference, or no statistical tests for the significance of the difference [16, 49, 57, 86, 87]. Only one study [21] adjusted for the clustering or reported estimates of differences between samples (odds ratio for insect-level positivity) together with intervals, using PoolTestR [98] for these calculations. A further study [41] determined a p-value for the prevalence difference by finding the largest confidence level for which the pairs of confidence intervals did not overlap, adjusting for pool testing but not clustering (Fig. 4).

**Fig 4:**
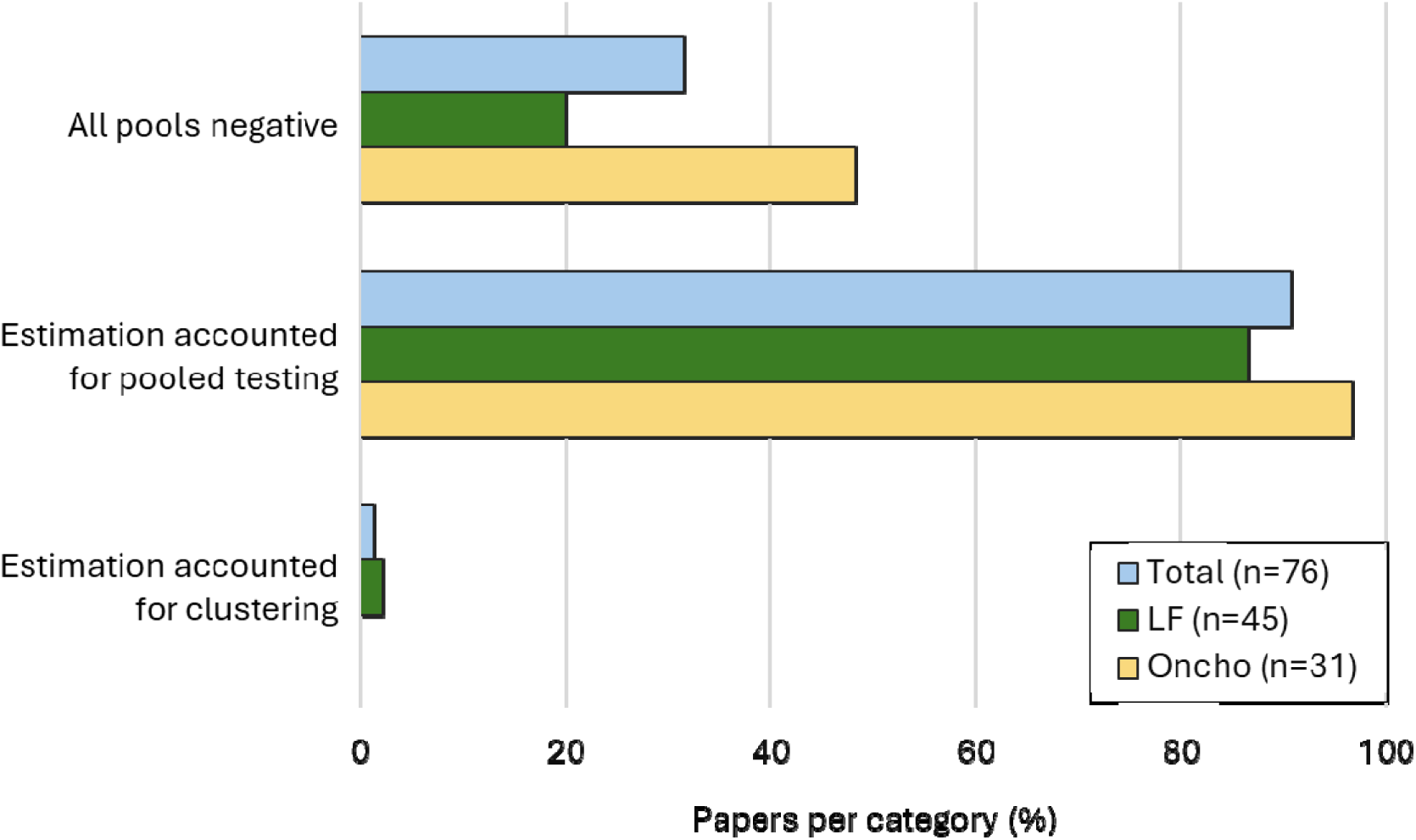
Frequency of key analysis choices and outcomes for the MX surveys in the included studies by disease and overall. Values are provided in Supplementary Table S3. Abbreviations LF: lymphatic filariasis; Oncho: onchocerciasis.

## Discussion

Our systematic review found that MX surveys have diverse objectives, designs and analysis methods. While it is beyond the scope of this review to make a detailed critique of the alignment between objectives, designs, and analysis for each of the 76 included studies, we highlight common misalignments between survey design and analysis that we have identified and suggest tools or resources that could be used to improve alignment.

While there was a near universal adoption of hierarchical cluster sampling designs, almost no studies adjusted for clustering in their analysis. The implications of this analytical omission are not trivial. Many studies aimed to compare their estimates (and confidence intervals) of prevalence to thresholds values, either thresholds required for the WHO certification of elimination (onchocerciasis) or provisional thresholds (LF). Failure to account for clustering in the analysis of data from cluster surveys may have led to artificially narrow confidence intervals [98] and therefore, undue confidence that prevalence was below a specified threshold. Similarly, other comparisons of prevalence, for example before and after interventions, could equally be in question due to a failure to account for clustering. The absence of software with the capability to easily adjust pool-tested data for clustering likely explains this major analytical omission. The only publicly available software with this capability, PoolTestR [98], was published by some of the authors of this review, in 2021 towards the end of the review period. Prior to this, the vast majority of studies reported using the PoolScreen software [96]. In the small number of studies where there was only a single sampling site [23] or only a single sampling site per study area [38], adjusting for clustering at sampling sites would not have been necessary; however, these sampling designs makes it difficult to generalise findings at the sample sites to the broader study population.

Only two studies indicated that they accounted for the loss of statistical power inherent to cluster and pool-testing designs by including design effects to increase their target sample sizes [21, 69]. However, these studies did not justify their choice of design effect on statistical grounds, so it is difficult to judge whether the additional sampling effort was sufficient. There was also generally insufficient justification of sampling strategies and pool sizes used in studies. Common design choices, such as the number of units in pools, appear to have been chosen primarily because others have made the same choice before, without any evidence to indicate that it was optimal for their study objectives based on statistical, laboratory, or practical grounds.

Pooling schemes varied substantially by studies. Many studies formed pools by combining insects collected from multiple locations. Though there are techniques using results from mixed pools to attempt to estimate prevalence for each population from which individual samples were collected [100] the available open-source software that can conduct these analyses [97, 101] cannot account for clustering at collection sites. Moreover, using pools from mixed locations inevitably leads to the loss of information even if analysed correctly.

Many studies fixed the pool size to be used across the survey, e.g. all pools contained exactly 20 insects. Studies that used a fixed pool size rarely stated what was done with remaining insects and suggests possible inefficiencies in the use of resources. If any remaining insects are not tested, this reduces the total sample size and discards potential information that could be gained by testing all insects. If sampling continues until a target number of insects are captured (e.g. nightly trapping until a quota is reached), then more sampling effort (e.g. trapping nights) will be dedicated towards collecting insects in the locations with lowest insect yields. In either case, this may be inefficient in settings where the highest cost component is sample collection. Though a detailed assessment of whether the pool sizes were appropriate in each of these studies would require the development of new statistical theory on optimal pool sizes in cluster surveys and is beyond the scope of this review, in some studies, the pools may have been too large as nearly all the pools were positive [21]. Similarly, though we cannot comment more generally on the optimal number of pools per site and this information was not provided in most studies, some studies included very few (often only one) pools per location [32, 83]. With only a single observation per location, it would not be possible to estimate the degree of location-level clustering of outcomes, which in turn would make it difficult to assess how estimates from samples could be generalised to unsampled locations.

In about one third of studies, none of the tested insects were positive for pathogen DNA. If one assumes (near) perfect sensitivity of the test, a negative pool implies that all the constituent insects would have also been negative if tested individually and therefore, statistical analysis does not require adjustment for testing in pools. However, in surveys attempting to validate elimination status of a disease by comparing prevalence estimates and their confidence intervals to a threshold, there is still a need to adjust for clustering within sample sites to ensure that widths of confidence intervals are not underestimated. None of the studies with all negative samples adjusted for clustering in their analysis, and the software used to adjust for clustering in MX studies [98] remains to be validated in such a setting. However, there is a fundamental difficulty in estimating the degree of clustering of infection from a dataset with no evidence of infection. If the degree of clustering of infection is estimated in a wide range of MX studies from around the world, these could be used as a prior to inform estimates of clustering (Bayesian paradigm) or as assumed values for computing confidence intervals (frequentist paradigm) when analysing data with no positives, or evaluating survey designs before sampling commences and the degree of clustering is unknown.

In many studies, data collected from multiple sites (e.g. households) across smaller geographical units (e.g. sentinel villages) were combined to estimate prevalence for a larger geographical area (e.g. region/state/focus); however, as many studies either did not state how the smaller geographical units were selected or selected them purposively [44, 49, 52, 59, 79, 87, 90], it is unclear whether data from the smaller geographical units could be validly combined to get unbiased estimates of prevalence in the larger geographical areas. Many of these studies used MX to evaluate elimination status after many rounds of MDA, in which case there was an obvious case to be made for selecting the highest risk locations for sampling. However, this approach is at odds with a threshold-based approach as currently required by the WHO for onchocerciasis, where an estimate of population prevalence (and therefore a population-representative survey) is required to compare to the target threshold.

Though there are published guidelines for the use of MX in onchocerciasis and LF [35, 91, 92, 94, 102], they do not and cannot include suggested designs that consider the resources and contextual constrains of each study or surveillance programme. Tools that enable MX practitioners to evaluate a wide range of MX designs and identify those best suited to their study objectives and constraints could fill this gap. Though there is an extensive literature and many publicly available software tools for selecting and evaluating the statistical properties (e.g. power, sample size calculations) suited for cluster surveys, there is no validated or widely accepted software which provides these tools for surveys using pooled testing.

There is an extensive literature and numerous software applications dedicated to the analysis of either pool-tested data or cluster data. However, little has been written about surveys that use pooling *and* cluster designs [103–105]; software that can analyse these surveys have only recently become available [98], and these tools still have gaps. For instance, many studies in this review compared prevalence between two or more samples, but did not quantify the differences, or conduct formal tests for the significance of these differences with methods that accounted for the cluster *and* pool-testing designs. A 2017 study highlighted the lack of suitable statistical tools for such analyses [20]. A subsequent study (2022) [21] used a regression model with the PoolTestR software [98] (2021) to make these comparisons; however, tools that simplify these comparisons may widen the use these types of analyses, important for evaluating interventions or confirming trends. LF and onchocerciasis are broadly acknowledged to be highly focal diseases (ref). However, none of the studies in the review used a spatial framework to design or analyse their data. Spatial sampling and analysis schemes can substantially reduce the sample size required to estimate population prevalence [106]. A geospatial modelling framework has been developed and applied for MX for tick-borne disease surveillance [107], but these models assume that all positive pools of vectors are retested to determine the infection status of individual vectors and are therefore would not be applicable to the vast majority of MX survey designs considered in this review. Applicable Bayesian geostatistical modelling approaches are possible with the PoolTestR software [98]; however, there are no published studies that demonstrate this kind of analysis with field data.

The final design and implementation of any MX survey will always be constrained by the available resources, and by the practical logistics of field work. Nevertheless, there has been an apparent tendency amongst researchers to neglect several important aspects of survey design unique to the hierarchal surveys and pooled data analysis that are commonly employed in MX surveillance for NTDs. The failure to consider and adjust for the implications of clustering on estimated prevalence is likely perpetuated by the paucity of examples in the literature which do so, and a lack of freely available and easy to use tools to facilitate the analysis. Such examples and tools are urgently needed in the MX space to improve the quality of the information being provided to inform major programmatic decisions on disease elimination.

## Data Availability

All data produced in the present study are available upon reasonable request to the authors

## Acknowledgment/Funding

This review was partially funded by a grant from the Australian Centre for Control and Elimination of Neglected Tropical Diseases, which was an Australian National Health and Medical Research Council (NHMRC; www.nhmrc.gov.au) Centre of Research Excellence Grant (number: 1153727). CLL was supported by an NHMRC Fellowship (number: 1193826). AM was supported by a Linkage grant (number: LP220100003) with funding from the Australian Research Council, and the Australian Department of Agriculture Fisheries and Forestry.

## Supplementary Materials

**Supplementary Table S1:** Counts (%) papers per category that address each of the five most common objectives, noting that papers can have multiple objectives.

| Objective | Onchocerciasis | LF | Overall |
| --- | --- | --- | --- |
| Post-MDA elimination determination | 27 (87) | 21 (47) | 48 (63) |
| Compare MX and human indicators | 18 (58) | 27 (60) | 45 (59) |
| Evaluate an intervention | 11 (35) | 8 (18) | 19 (25) |
| Compare collection methods | 0 (0) | 13 (29) | 13 (17) |
| Compare MX lab techniques | 5 (16) | 13 (29) | 18 (24) |

**Abbreviations** LF: lymphatic filariasis; MX: molecular xenomonitoring

**Supplementary Table S2:** Counts (%) of key design choices and justifications for the MX surveys in the included studies.

| Design choice | Onchocerciasis | LF | Overall |
| --- | --- | --- | --- |
| Hierarchical sampling frame | 29 (94) | 43 (96) | 72 (95) |
| Justified sample sizes | 9 (29) | 7 (16) | 16 (21) |
| Justified pool size | 2 (6) | 3 (7) | 5 (7) |
| Considered design effects in sample size calculations | 0 (0) | 2 (4) | 2 (3) |
| Same number of units in (nearly) all pools | 13 (42) | 17 (38) | 30 (39) |

**Abbreviations** LF: lymphatic filariasis

**Supplementary Table S3:** Counts (%) of key analysis choices and outcomes for the MX surveys in the included studies.

| Analysis choice or outcome | Onchocerciasis | LF | Overall |
| --- | --- | --- | --- |
| All pools negative | 15 (48) | 9 (20) | 24 (32) |
| Estimation accounted for pooled testing | 30 (97) | 39 (87) | 69 (91) |
| Estimation accounted for clustering | 0 (0) | 1 (2) | 1 (1) |

**Abbreviations** LF: lymphatic filariasis

